# Persistent financial adversity and cognitive ageing: A lifecourse investigation

**DOI:** 10.1101/2024.10.20.24315831

**Authors:** Yiwen Liu, Jacques Wels, Sarah-Naomi James, Sarah E Keuss, Jane Maddock, Thomas D Parker, Jean Stafford, Jonathan M Schott, Marcus Richards, Praveetha Patalay

**Author notes:** **Corresponding author**: Dr Jacques Wels, MRC Unit for Lifelong Health & Ageing, University College London, London, UK, WC1E 7HB.

## Abstract

**Background:** Little is known about the effect of persistent financial adversity across adulthood on cognitive ageing, and whether these impacts vary based on sex, childhood socioeconomic circumstances (SEC) and genetic risk.

**Methods:** Using data from the 1946 Birth cohort study (N=2,759), with extensive data spanning over 70 years, as well as an embedded neuroimaging study (Insight46, *N*=356-468), we examined the prospective association between financial adversity (low household income, financial hardships; 26-53 years) and cognitive ageing (cognitive performance at 53 years; decline between 53-69 years, modelled using latent growth curve model; neuroimaging measures of brain health between 69-74 years), and the moderating role of sex, childhood SEC and APOE-4. Covariates included sex at birth, childhood SEC, childhood cognition (8 years), symptoms of depression and anxiety (13-15 years), and educational attainment (26 years).

**Findings:** Increased exposure to low household income as well as financial hardships was associated with lower processing speed (SE: -0.07 [95% CI: -0.13, -0.02], -0.05 [-0.11, - 0.00], respectively) and verbal memory at age 53 (-0.16 [-0.21, -0.11], -0.10 [-0.15, -0.05] respectively). Increased exposure to financial adversity was also associated with slower verbal memory decline from 53 to 69 years, due to already lower baseline scores at 53 years. Persistent financial adversity was associated with greater ventricular volume at 69-71 years, and stronger associations between financial adversity and brain atrophy were found for males, those with lower childhood SEC, and APOE-4 carriers. APOE-4 carriers in particular were consistently more vulnerable to the effect of persistent financial adversity on brain atrophy.

**Interpretation:** Persistent exposure to financial adversity influences cognitive performance by midlife and later-life brain atrophy, with impacts being larger for males, disadvantaged childhood SEC and individuals with greater genetic risk. These highlight the potential role of poverty reduction efforts in working-age adults for preventing dementia and promoting cognitive health in an ageing population.

**Funding:** This study is funded by the UK Medical Research Council which provides core funding for the NSHD (MC_UU_00019/1 and MC_UU_00019/3). Insight 46 is funded by grants from Alzheimer’s Research UK (ARUK-PG2014-1946, ARUK-PG2017-1946: JMS, MR), Alzheimer’s Association (SG-666374-UK BIRTH COHORT: JMS), the Medical Research Council Dementias Platform UK (CSUB19166: JMS, MR), The Wolfson Foundation (PR/ylr/18575: JMS), The Medical Research Council (MC-UU-12019/1: Kuh and MC-UU-12019/3: MR), Selfridges Group Foundation (22/3/18: JMS), and Brain Research Trust (UCC14191: JMS). JS, JM and PP are also supported by funding from Alzheimer’s Society (Ref:469). TDP is supported by a NIHR clinical lectureship. JW is funded by the Belgian National Scientific Fund (FNRS) Research Associate Fellowship (CQ) no. 40010931.

## Introduction

Dementia is one of the leading contributors to the global burden of disease,^1,2^ and the rates are estimated to triple to 150 million over the next three decades.^3^ There is a need to better understand how modifiable environmental exposures may be associated with cognitive ageing.^4^ The association between childhood and adulthood socioeconomic circumstances (SEC) and cognitive functioning has been shown at different stages of the lifecourse.^5–8^ Financial adversity – commonly defined as having a lack of money or resources to provide basic household necessities^9^ – is also associated with cognitive impairment and decline.^5,10–14^ Although there is evidence for lifecourse and intergenerational transmission of poverty^15,16^ most studies to date use a snapshot measure of financial adversity that does not capture the persistence of these experiences.

Financial adversity has been assessed using either objective (e.g., household income) or subjective (e.g., perceived financial hardships such as inability to pay bills) indicators, with only one study identified having examined both in relation to cognitive function in midlife.^17^ Although cognitive ageing is often assessed as cognitive impairments or individual rates of decline using neuropsychological measures, neuroimaging studies can also index brain health markers that are associated with cognitive decline and dementia.^18–21^ The association between persistent experiences of financial adversity and cognitive ageing may also be moderated by other biological or environmental factors, including sex, genetic risk, or childhood SEC, which remains to be investigated.

Testing the longitudinal association between persistent financial adversity and cognitive ageing requires the use of studies that have followed participants through multiple stages of development, such as the 1946 NSHD (National Survey of Health and Development) cohort. It is one of the world’s oldest continuously running birth cohort studies with more than 70 years of follow-up since birth,^22^ with repeated measures of financial adversity and cognitive function, and an embedded neuroimaging sub-study. Furthermore, extensive information collected in childhood allows controlling for potential confounder variables (particularly childhood cognitive ability) and enables testing of potential moderating effects of sex, genetic risk and childhood SEC.^23,24^

The overall aim of this study was to examine the effect of persistent financial adversity across adulthood on indicators of cognitive ageing including cognitive decline and markers of brain health. We specifically investigated three questions: (1) what is the association between persistent financial adversity and cognitive function from mid-to-late adulthood; (2) what is the association between persistent financial adversity and markers of brain health; and (3) is there a moderating effect of sex, childhood SEC, and genetic risk (APOE-ε4)?

## Methods

### Pre-registration with OSF

(https://doi.org/10.17605/OSF.IO/6Y9NV).

### Sample

The MRC 1946 NSHD (*N*=5,362) is a national British birth cohort of people born in one week of March 1946 (http://www.nshd.mrc.ac.uk/nshd).^22^ The main analytical sample included participants still alive at 69 years and with available cognitive assessments at age 53, 63, or 69 (*N*=2,759) (see supplement, Fig. S1). Ethical approval has been obtained from the Greater Manchester Local Research Ethics Committee and the Scotland Research Ethics Committee, and participants provided written informed consent at each data collection.

502 NSHD participants (mean [SD] age 70.7 [0.7] years) were recruited to the Insight 46 sub-study for β-amyloid (Aβ) positron emission tomography (PET) and MR imaging based on previously published criteria.^25,26^ Of those who took part, 471 completed a brain scan at baseline and 369 completed a follow-up scan (mean [SD] scan interval 2.4 [0.2] years) (supplement, Fig. S1).

## Measures

### Financial adversity (26-53 years)

#### Household income

Total household income was assessed at 26, 43 and 53 years, and was comparable to the population average at the time (see supplement for details). A binary variable was derived at each time point by grouping those with household income in the bottom 20% of the analytical sample^27^ into the low-income category. These were then summed across the three time points, providing a count of experiences of low household income (0 to 3) and further grouped into none (0), intermittent (1) or persistent experience (≥2).

#### Financial hardships

Financial hardships were assessed at 36, 43 and 53 years (see supplement). A binary variable was derived at each time point and summed across the three time points, providing a count of experiences of financial hardships (0 to 3). This was further grouped into none (0), intermittent (1) or persistent experience (≥2).

### Cognitive function (53, 63, 69 years)

Processing speed and verbal memory were assessed using the same assessments at 53, 63 and 69 years, with scores standardised across all ages.

#### Processing speed

This was assessed using a timed-letter search task where participants had to cross out target letters “P” and “W” embedded among non-target letters within 1 minute. A score represented the position reached at the end of the trial (max = 600).

#### Verbal memory

This was a word learning task, where participants were shown a list of 15 words over 3 administrations and were asked to write down as many as they could remember at the end of each. A score indicated the number of words correctly recalled across all administrations (max = 45).

### Brain health

The full neuroimaging protocol for Insight 46 has been published.^26^ Structural MRI sequences first underwent correction for gradient non-linearity,^28^ brain-masked N4-bias correction,^29^ and visual inspection of image quality. The following measures were included (detailed description in supplement): (1) baseline (69-71 years) global white matter hyperintensity volume (WMHV); (2) baseline β-amyloid (Aβ) burden (standardized uptake value ratios [SUVR]) and status (positive/negative); (3) baseline whole brain, ventricular and total hippocampal volume; and (4) longitudinal changes in whole brain, ventricular, and hippocampal volume between baseline and repeat MRI (71-74yrs), calculated using the Boundary Shift Integral (BSI)^30–32^ described elsewhere.^33^

### Covariates

Covariates included: (i) sex at birth; (ii) childhood SEC (paternal occupational status grouped into non-manual/skilled vs manual/unskilled/unemployed); (iii) childhood cognition (8 years, summary measure of 4 tests from the National Foundation for Educational Research); (iv) symptoms of depression and anxiety (13-15 years), rated by teachers and grouped into absent, mild or severe; and (v) educational attainment (26 years), grouped into ordinary (‘O’)-level or below (equivalent to 11 years of education) vs. advanced (‘A’) level or above, with the latter as the reference. Total intracranial volume, calculated using Statistical Parametric Mapping (SPM) 12,^34^ was included as a covariate in models with volume outcomes, to adjust for head size.^23^

### Moderators

Sex, childhood SEC and APOE-ε4 – which was genotyped using two SNPs (rs439358 and rs7412) at 53 years and grouped into non-carrier vs carrier (homozygous or heterozygous). .

### Statistical analysis

All statistical analysis was performed in R (version 3.6.2).

### Cognitive function outcomes

A latent growth curve (LGC) model was used to examine changes in cognitive function over time.^35^ Latent variables were generated for the intercept (baseline performance at 53 years) and slope (change from 53 to 69 years, with lower scores indicating greater decline). Time in the model was centred on the first assessment period (53 years) and converted to decades. Statistical formula can be found in supplement. Full information maximum likelihood (FIML) was used to estimate cognition intercept and slope within the LGC model.

After deriving cognition intercept and slope from LGC models, linear regressions were estimated to examine their association with persistent financial adversity (low household income and financial hardships, which were examined in separate models and treated as ordered variables to examine linear trends). These models were adjusted for all covariates.

To test for non-linearity in dose-response, low household income and financial hardships were additionally treated as categorical variables in regression models, and quantile regression analysis investigated whether the effect of persistent financial adversity was similar across different quantiles (25^th^ and 75^th^) of cognitive decline (see supplement).

To examine effect modification, we included interaction terms between indicators of financial adversity and each of the modifiers on cognition intercept and slope in separate models. Effect modifications with p-value <0.10 were visualised using predicted means from the model.

### Brain health outcomes

The association between persistent financial adversity and markers of brain health were estimated using logistic regression (for Aβ status) and linear regression (all other outcomes). WMHV was log-transformed due to its positively skewed distribution. Longitudinal change in volumetric measures (BSI) were further adjusted for time interval between MRI scans by dividing each BSI metric by time (years) between scans (to represent change per year).

Three models were fitted for each marker of brain health: the first examined the main effect of financial adversity, adjusted for sex and intracranial volume (for volumetric measures). The second examined the main effect of each moderator, adjusted for all covariates. The third examined the interaction between financial adversity and each moderator, adjusting for all covariates. Significant interactions were plotted using predicted means from the model.

### Missing data

To account for attrition, inverse probability weighting (see supplement) was used to calculate non-response weights for the analytic sample compared to the whole NSHD sample. Within the analytic sample, multivariable imputation by chained equations (MICE) was used (20 imputations) for other study variables (see supplement).^36,37^

### Additional and sensitivity analyses

Additional interactions were tested between the level of financial adversity (persistent or intermittent) associated with cognitive decline and each moderator, and between low household income and financial hardships (if both were associated with any outcome of interest). Four sensitivity analyses were also included to check the robustness of findings; primary analyses were repeated: (1) without sample weightings; (2) within Insight 46 participants only; (3) using a household size adjusted low income variable; (4) performing multiple imputation prior to performing LGC analysis.

## Results

Of 2,759 participants included in the main analytical sample, 16% and 12% reported persistent low household income and financial hardships. The majority (73.6%) of those who never experienced low household income also never experienced financial hardships, and vice versa (66.9%) (Table 1).

**Table 1.** Sample characteristics.

|  | **Overall** | **Low household income** |  |  | **Financial hardships** |  |  |
| --- | --- | --- | --- | --- | --- | --- | --- |
|  |  | *None* | *Intermittent* | *Persistent* | *None* | *Intermittent* | *Persistent* |
|  | 2759 | 1574 | 733 | 426 | 1727 | 689 | 313 |
|  | N (% of total) | N (% of each group) |  |  | N (% of each group) |  |  |
| **Low household income** |  |  |  |  |  |  |  |
| None | 1574 (57.6) | - | - | - | 1150 (66.9) | 322 (47.0) | 90 (28.8) |
| Intermittent | 733 (26.8) | - | - | - | 412 (24.0) | 223 (32.6) | 93 (29.7) |
| Persistent | 426 (15.6) | - | - | - | 156 (9.1) | 140 (20.4) | 130 (41.5) |
| **Financial hardships** |  |  |  |  |  |  |  |
| None | 1727 (63.3) | 1150 (73.6) | 412 (56.6) | 156 (36.6) | - | - | - |
| Intermittent | 689 (25.2) | 322 (20.6) | 223 (30.6) | 140 (32.9) | - | - | - |
| Persistent | 313 (11.5) | 90 (5.8) | 93 (12.8) | 130 (30.5) | - | - | - |
| **Sex [Female]** | 1416 (51.3) | 743 (47.2) | 408 (55.7) | 255 (59.9) | 913 (52.9) | 333 (48.3) | 156 (49.8) |
| **Childhood SEC [unskilled/unemployed]** | 241 (11.0) | 104 (8.5) | 75 (12.7) | 58 (16.5) | 135 (9.8) | 61 (11.2) | 44 (18.3) |
| **Childhood symptoms of depression and anxiety** |  |  |  |  |  |  |  |
| Absent | 1260 (51.2) | 769 (55.1) | 319 (48.6) | 163 (41.3) | 802 (51.5) | 318 (53.0) | 129 (45.3) |
| Mild | 936 (38.0) | 511 (36.6) | 257 (39.2) | 164 (41.5) | 596 (38.3) | 218 (36.3) | 116 (40.7) |
| Severe | 265 (10.8) | 116 (8.3) | 80 (12.2) | 68 (17.2) | 158 (10.2) | 64 (10.7) | 40 (14.0) |
| **Educational attainment [O-level or below]** | 1655 (63.2) | 819 (54.7) | 488 (70.4) | 336 (81.8) | 982 (59.4) | 442 (68.5) | 218 (73.9) |
| **APOE-ε4 status [Carrier]** | 702 (30.1) | 413 (31.0) | 186 (29.3) | 102 (28.7) | 452 (30.9) | 173 (29.0) | 77 (28.7) |
| **Processing speed 53 years (mean (SD))** | 283.61 (75.35) | 287.62 (73.55) | 283.50 (76.09) | 269.09 (78.41) | 286.61 (74.35) | 282.80 (77.66) | 269.23 (74.19) |
| **Processing speed 63 years (mean (SD))** | 267.12 (71.33) | 271.39 (70.29) | 266.79 (73.47) | 249.46 (69.73) | 268.05 (71.47) | 268.31 (70.84) | 255.76 (71.03) |
| **Processing speed 69 years (mean (SD))** | 262.41 (74.18) | 263.94 (70.54) | 264.61 (80.39) | 250.39 (75.96) | 264.28 (73.49) | 261.94 (75.60) | 253.07 (73.95) |
| **Verbal memory 53 years (mean (SD))** | 24.13 (6.25) | 25.19 (6.01) | 23.45 (6.42) | 21.55 (5.86) | 24.72 (6.14) | 23.37 (6.26) | 22.54 (6.37) |
| **Verbal memory 63 years (mean (SD))** | 24.28 (6.10) | 25.15 (6.05) | 23.72 (5.99) | 21.54 (5.65) | 24.83 (6.06) | 23.46 (6.09) | 22.84 (6.02) |
| **Verbal memory 69 years (mean (SD))** | 22.16 (6.07) | 23.00 (5.97) | 21.29 (6.02) | 20.16 (5.87) | 22.66 (5.92) | 21.57 (6.06) | 20.68 (6.55) |

### Cognitive function outcomes

Increased experience of financial adversity showed dose-response associations with processing speed (low household income: SE = -0.13, 95%CI [-0.18, -0.08]; financial hardships: -0.09 [-0.14, -0.04]) and verbal memory (low household income: -0.34 [-0.40, - 0.29]; financial hardships: -0.24 [-0.30, -0.17]) at 53 years. These effect sizes were somewhat attenuated after adjusting for covariates (supplement Table S1).

There was overall weak evidence for an association between financial adversity and processing speed decline, but a small dose-response effect was found on verbal memory decline, with increased exposure associated with slower decline (low household income: 0.02 [0.01, 0.02]; financial hardships: 0.01 [0.01, 0.01]) (supplement Table S1). Further testing of non-linearity showed that only persistent experience of financial adversity (rather than intermittent) was associated with slower verbal memory decline (supplement Table S2, S3). This may be explained by those with lower baseline cognitive performance also showing a slower rate of decline over time. To illustrate this, we stratified the sample based on baseline cognition, and those scoring 1 SD or below at 53 years showed a slower rate of decline particularly if they also persistently experienced financial adversity (Fig.1; supplement Fig S3).

**Fig. 1.**
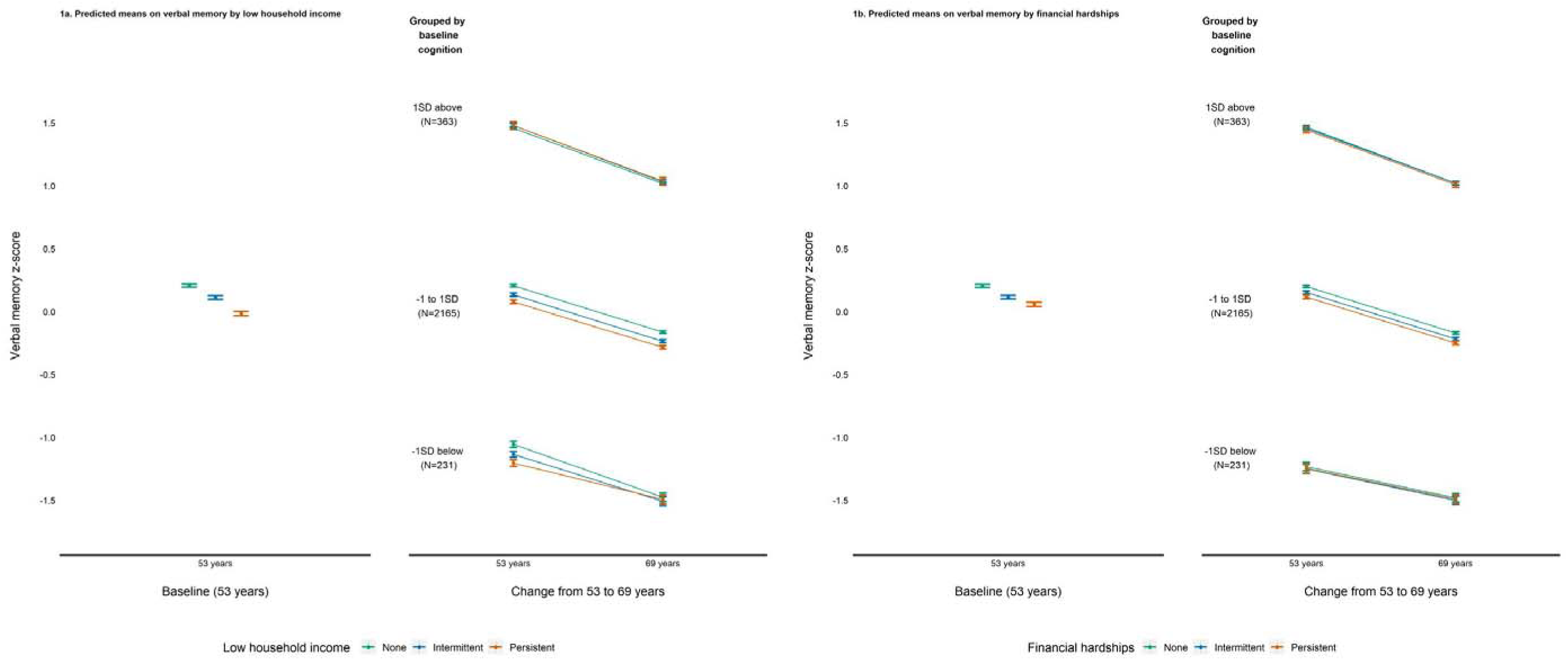
Predicted means on verbal memory at baseline (53 years) and decline (stratified on cognition at baseline) by financial adversity exposure.

Sex moderated the effect of low household income on processing speed at baseline (53 years) (p = 0.009), and the effect of financial hardships on processing speed decline (p = 0.075) (supplement Table S4). Post-hoc analyses showed that, compared to females, males who persistently experienced low household income scored lower on processing speed at 53 years, but showed slower decline when persistently experiencing financial hardships (Fig. 2). No other evidence of interactions was found for childhood SEC or genetic risk.

**Fig. 2.**
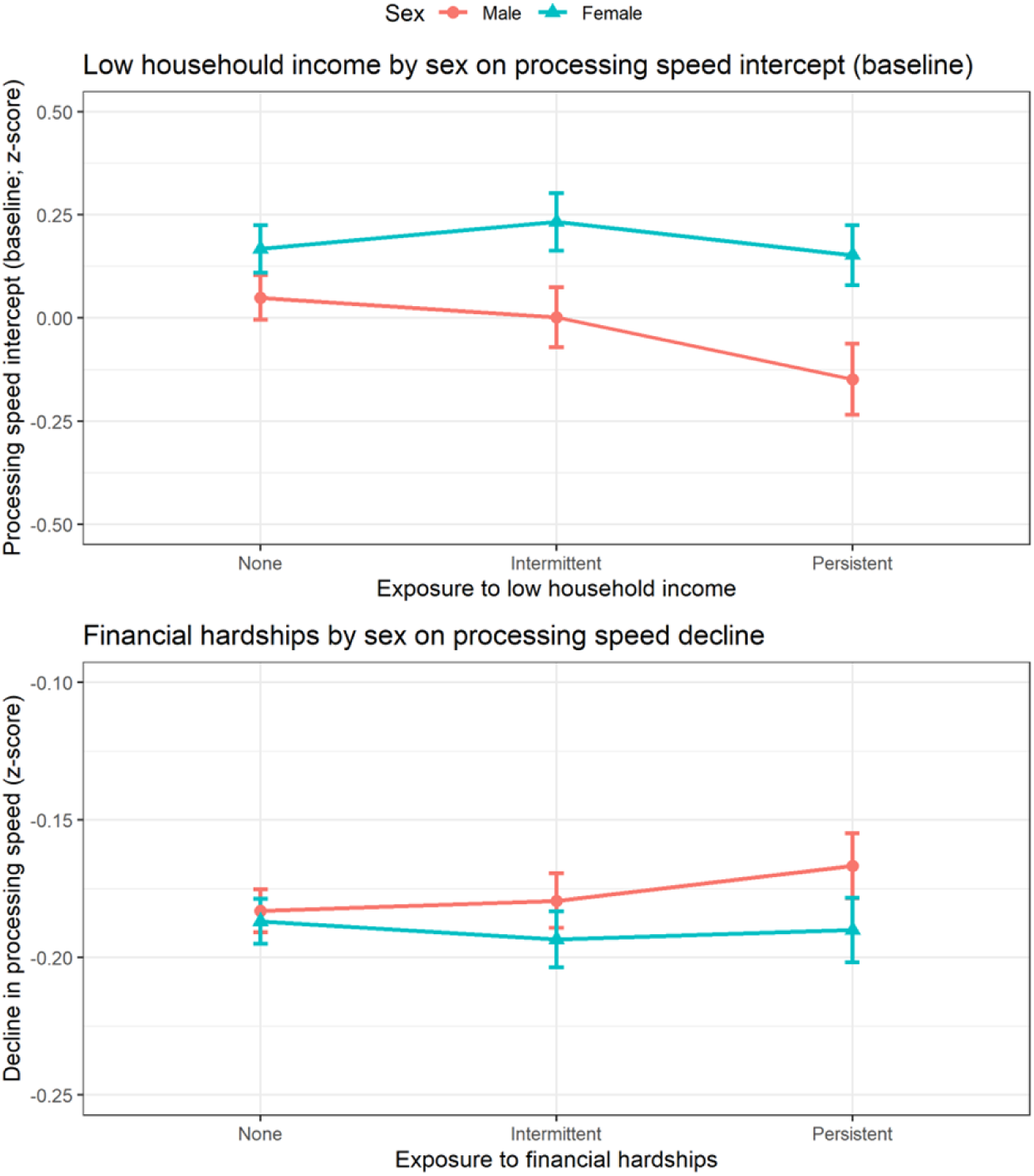
Predicted means on processing speed at baseline (53 years) and decline (lower scores representing faster decline) by financial adversity exposure and sex.

### Brain health outcomes

Increased low household income was associated with larger ventricular volume at 69-71 years (b = 4.67 [1.01, 8.32]). No other baseline associations were found (supplement Table S5). An interaction was found between financial hardships and APOE-4 on Aβ burden (SUVR) (p = 0.099), with carriers persistently experiencing financial hardships showing the highest Aβ burden (Fig. 3g; supplement Table S5). An additional sex interaction was found with financial hardships on baseline ventricular volume (p = 0.046), with larger volume seen in males who persistently experienced financial hardships compared to females (Fig. 2e; supplement Table S5)

No direct associations were found between financial adversity and longitudinal changes in volumetric measures. However, several interactions were identified (supplement Table S6). First, financial hardships and sex interacted on total brain volume atrophy (p = 0.003) and ventricular expansion (p = 0.005), with males who persistently experienced financial hardships showed faster brain atrophy and ventricular expansion compared to females (Fig. 2c-d). Second, low household income and childhood SEC interacted on total hippocampal volume atrophy (p = 0.033) and ventricular expansion (p = 0.031), where those with disadvantaged childhood SEC who persistently experienced financial hardships across adulthood showed faster hippocampal atrophy and ventricular expansion compared to those with advantaged childhood SEC (Fig. 2a-b). Finally, APOE-4 status showed both a main effect, as well as modified the effect of low household income and financial hardships on all volumetric changes (supplement Table S6). Carriers of APOE-4 who persistently experienced low household income or financial hardships showed faster total brain and hippocampal volume atrophy, as well as increased ventricular expansion, compared to non-carriers. (Fig. 3a-f).

**Fig.2.**
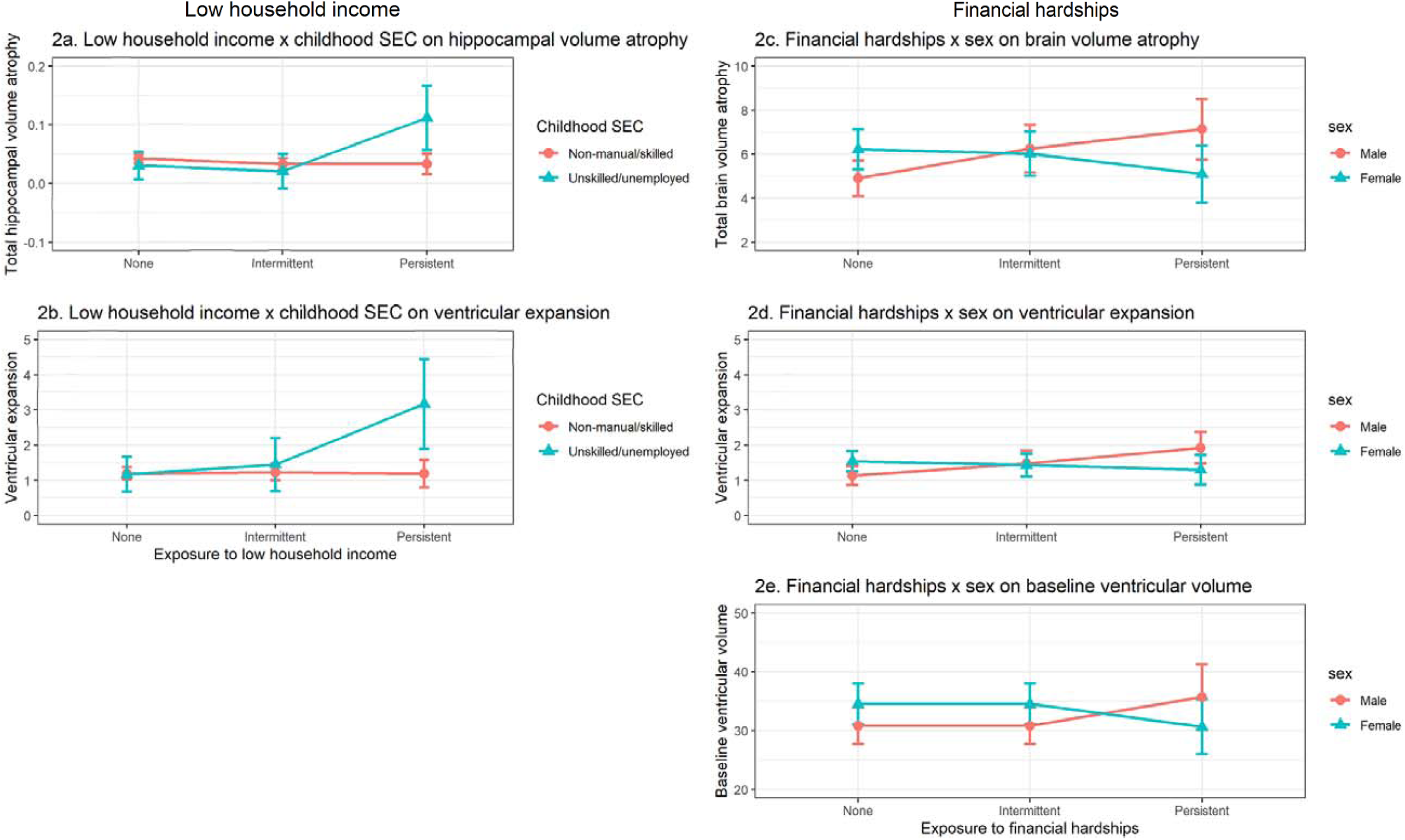
Predicted means on longitudinal measures of brain atrophy and baseline ventricular volume by financial adversity exposure and sociodemographic (sex, SEC) variables.

**Fig. 3.**
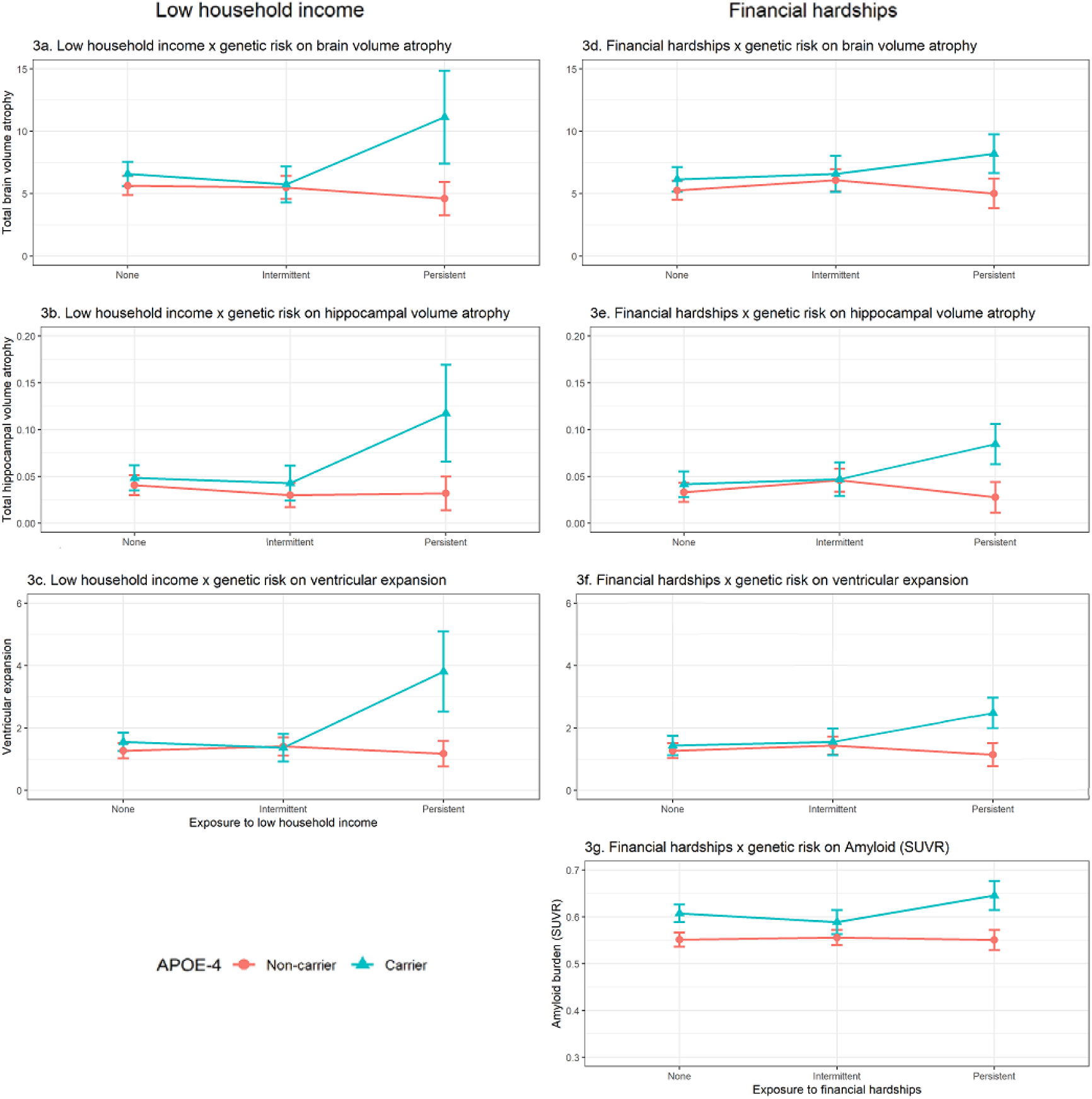
Predicted means on longitudinal measures of brain atrophy and baseline amyloid (Aβ) burden by financial adversity exposure and APOE-4 status.

### Additional and sensitivity analyses

No evidence of interactions was found for planned additional analyses (supplement Table S7, S8), and sensitivity analyses yielded similar effect sizes (outputs can be found in the supplement Tables S9-S15, Figures S4, S5).

## Discussion

The aim of this study was to examine the effect of persistent financial adversity on cognitive ageing. Findings add to the existing evidence base in at least 4 ways: first, we investigated the persistence of both objective (low household income) and subjective (financial hardships) measures of financial adversity over many decades in the same individuals, and second, focused on cognitive decline over almost 2 decades rather than a single measure at baseline. The findings that increased experience of both indicators of financial adversity were associated with lower scores on processing speed and verbal memory at 53 years, but a slower rate of decline for verbal memory between 53 and 69 years, suggest substantial cognitive losses already present at midlife. One mechanism proposed for the role of financial adversity in cognitive impairment is that financial adversity may increase cognitive load, thus reducing the available cognitive bandwidth involved in processes such as attention and decision making.^11,38–40^ Persistent experience of financial adversity, therefore, may place chronic stress on systems involved in cognitive processing and lead to cognitive impairments over time. Third, we included neuroimaging data across two timepoints and found a dose-response association between low household income and increased ventricular volume at 69-71 years. Fourth, we examined interactions with sex, childhood SEC and APOE-4, which all moderated the effect of financial adversity on cognitive ageing. APOE-4 in particular modified all associations with brain atrophy, such that carriers who persistently experienced low household income or financial hardships showed the highest rates of brain atrophy.

Sex also moderated the effect of financial adversity on processing speed, with males who persistently experienced financial adversity showing worse performance at 53 years^41^ and further showed a slower rate of decline over time compared to females, consistent with our proposal that those with lower cognitive performance at baseline show less decline over time. It has been suggested that males may be more vulnerable to the effect of financial adversity and low education on physical health outcomes,^42,43^ and findings here suggest that they may also be more vulnerable on cognitive outcomes. This may also be explained by males in the current cohort occupying more traditionally gendered roles compared to females, and therefore may be more adversely affected by experiences of financial adversity compared to females.

A dose-response effect was also found between financial adversity and brain health, with increased experience of low household income associated with a 4.7ml increase in ventricular volume at 69-71 years. In addition, there was evidence for multiple vulnerabilities to brain atrophy. Males appeared more vulnerable to the effect of persistent financial hardships on larger ventricular volumes and increased ventricular expansion, as well as increased total brain volume atrophy compared to females. Those with disadvantaged childhood SEC also appeared more vulnerable to the effect of persistent low household income on both hippocampal volume atrophy and ventricular expansion, extending previous findings.^44,45^ APOE-4 carriers who were also persistently experiencing financial adversity showed higher rates of Aβ deposition (SUVRs), increased total brain and hippocampal atrophy, as well as ventricular expansion, highlighting a multiplicative effect where those with greater genetic risk are particularly vulnerable to the effects of persistent financial adversity.

Hence the impact of persistent financial adversity on markers of brain health may be more complex depending on other co-existing factors such as sex, childhood SEC and genetic risk. APOE-4 has been shown previously to interact with educational attainment in its association with cognitive impairments and dementia risk, where higher education may protect against genetic susceptibility conferred by the APOE-4 allele.^46–49^ Similarly, our findings consistently showed that the association between APOE-4 and increased brain atrophy was only evident for those who persistently experienced financial adversity. As APOE-4 is hypothesised to act primarily via Aβ deposition^50^ – an early pathological feature of Alzheimer’s disease^51–53^ – the additional interaction found between financial hardships and APOE-4 on Aβ burden in the current study further suggest that environmental exposure may exacerbate the effect of APOE-4 on dementia risk. One potential mechanism for how APOE-4 may moderate the effect of persistent financial adversity is that persistent adversity may reduce brain and cognitive reserve, which, when combined with genetic susceptibility to pathology, may render systems more vulnerable and decrease the threshold for clinical symptoms.^46,54^ Reducing the frequency of experiencing financial adversity could therefore improve cognitive performance and help protect against the expression of genetic vulnerability to brain atrophy and subsequent risk of dementia.

Some limitations of the study include selective attrition bias, with those who dropped out being more likely to have lower cognitive abilities and disadvantaged SEC,^22^ particularly within Insight 46, which is a healthier and more socially advantaged cohort compared to the general population.^25^ This may lead to an underestimation of the effect of financial adversity on cognitive ageing. There is also a lack of ethnic diversity in this cohort, and further research is needed using more ethnically representative populations. Another limitation was the type of data collected for household income, which was a categorical variable indicating various income brackets rather than raw income values and not adjusted for household size; potentially reducing the precision of the low income variables derived.^55^ Other pathways from financial adversity to markers of brain health were also not investigated in this study, with one potential candidate being cerebrovascular dysfunction, which may precede neurodegenerative changes in the brain and cognitive decline.^56,57^

## Conclusion

Persistent financial adversity is associated with reduced cognitive function in midlife and subsequently slower decline into older age, highlighting substantial cognitive losses already present in midlife. Males, those with disadvantaged childhood SEC and greater genetic risk (APOE-4) were also more susceptible to the effect of persistent financial adversity on brain atrophy. Given the current climate of cost-of-living crisis where a record number of households are reporting financial adversity,^58^ these findings further illustrate the importance of supporting vulnerable households to prevent financial adversity from becoming chronic, which could also help protect against age-related cognitive impairments and disorders such as dementia.

## Supporting information

See supplement

## Data Availability

All data produced in the present study are available upon reasonable request to the University College London MRC unit for lifelong health and ageing (LHA).

## Acknowledgements

We are grateful to all study members who took part in the NSHD as well as Insight 46, and to the NSHD and Insight 46 study team members who helped to collect the data over the last seven decades. We are also grateful to the radiographers and nuclear medicine physicians at the UCL Institute of Nuclear Medicine, and to the staff at the Leonard Wolfson Experimental Neurology Centre at UCL.

## Author contributions

Conceptualisation: PP, all authors

Data curation and resources: SK, TP, JMS, MR

Funding acquisition and project administration: JMS, PP, MR

Methodology: YL, JW, S-NJ, JM, JS, PP

Formal analysis, visualisations: YL

Analysis verification: JW

Writing – first draft: YL

Writing – revision and final draft: all authors

## Data availability

Data will be made available to researchers upon request to the NSHD Data Sharing Committee. For more information see: http://www.nshd.mrc.ac.uk/data.

## Competing interest

The authors declare no competing interests.

## Notes

### Competing Interest Statement

The authors have declared no competing interest.

### Author Declarations

We used data from the MRC 1946 NSHD,a national British birth cohort. Ethical approval has been obtained from the Greater Manchester Local Research Ethics Committee and the Scotland Research Ethics Committee, and participants provided written informed consent at each data collection.

