## Supplementary material for "Persistent financial adversity and cognitive ageing: A lifecourse investigation": See supplement

**[Supplementary materials]**

### **Methods**

#### ***Sample selection and characteristics***

Sample selection for NSHD and Insight 46 can be found in Fig. S1. Those who continued participation in NSHD were more likely to be in non-manual occupations, have higher educational attainment and childhood cognitive ability compared to non-responders.^1^ Participants recruited to Insight 46 were more likely to be from higher SEC, with better mental health and self-rated health, and were less likely to be obese or a current smoker compared to those who were invited but did not attend the clinic.^2^

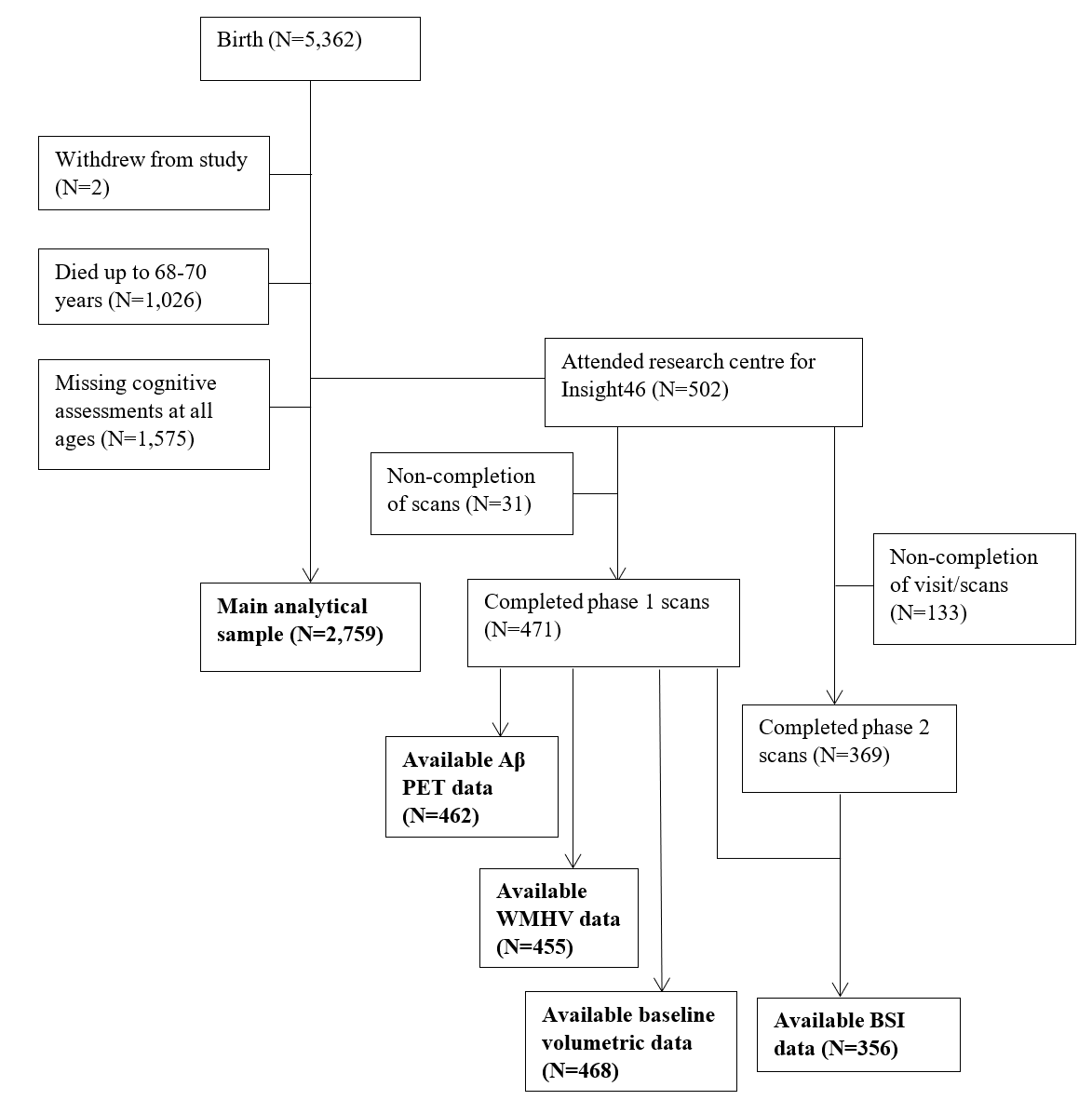

**Fig. S1**. Sample flow chart.

*Abbreviations: Aβ = β-amyloid; PET = positron emission tomography; WMHV: white matter hyperintensity volume; BSI = Boundary Shift Integral (assessing changes in brain volumes between baseline and follow-up).*

#### ***Measures***

###### Household income

Household income included regular payments as well as any benefits or pensions received, after deductions for tax and national insurance. This was reported as a monthly figure at age 26 and as an annual figure at 43 and 53 years. Participants were presented with cards indicating different income brackets and were asked to choose the card that best matched their own. Income brackets ranged from <£40 to ≥£200 monthly at 26 years (in £20 increments, i.e., £40-£59, £60-£79), and from <£2,000 to ≥£50,000 annually (in £1000 increments up to £12,000, followed by £12,000-£14,999, and in £5000 increments thereafter) at 43 and 53 years. The median household income in this cohort was £120-129 monthly at 26 years (UK in 1972: £133.6 for males, £74.4 for females), £11,000-£11,999 annually at 43 years (UK in 1989: £13,109), and £20,000-£24,999 annually at 53 years (UK in 1999-2000: £22,004).^3,4^

###### Financial hardships

Financial hardships were assessed at 36, 43 and 53 years. Across all three time points, participants were asked (i) how they managed on their current income (hard to manage, manage fairly well, or manage very well), and (ii) whether they had to go without things in the last 12 months due to a shortage of money (yes or no). An additional item on whether participants (iii) had trouble paying for bills in the last 12 months (never, sometimes, or often) was included at 43 and 53 years only.

###### Brain health

(1) Baseline white matter hyperintensity volume (WMHV): Generated by applying Bayesian Model Selection (BaMoS), an unsupervised automated algorithm ^5^ to T1 and FLAIR MRI. A global WMHV measure was generated which included subcortical grey matter but not infratentorial regions.

(2) Baseline Aβ status and burden: Baseline Aβ PET data were acquired after intravenous injection of the 18F Aβ PET ligand florbetapir. Aβ PET data from the 10-minute period, approximately 50 minutes post-injection, was processed including pseudo-CT attenuation correction.^6^ Baseline Aβ global SUVRs (standardized uptake value ratios) were generated from a composite cortical region of interest,^7^ and an eroded subcortical white matter reference region. Aβ status (positive/negative) was determined by applying gaussian mixture modelling to global SUVRs, and taking the 99^th^ percentile of the lower gaussian as the cut-point for positivity (SUVR>0.6104).^8^

(3) Baseline whole brain, ventricular and hippocampal volumes: Baseline whole brain and hippocampal volumes were automatically segmented from T1 MRI using Multi-Atlas Propagation and Segmentation (MAPS) and Similarity and Truth Estimation for Propagated Segmentation (STEPS) respectively.^9,10^ Images were visually checked for segmentation errors and manually edited if needed. Baseline ventricular volumes were determined from T1 MRI using a semi-automated technique.^11^

(4) Longitudinal changes in whole brain, ventricular and total hippocampal volume: Changes between baseline (age 69-71) and repeat T1 MRI (age 71-74) were calculated using the Boundary Shift Integral (BSI)^12–14^ as described elsewhere.^15^ All scan-pairs were reviewed to check that image quality was consistent between time-points.

#### ***Analysis and statistical equations***

***Primary analyses***

*Change in cognitive functioning over time*

Change in cognitive functioning between 53 and 69 years will be examined using latent growth curve (LGC) analysis. The formula for LGC is shown below:^16^

$$y_{i}=a_{1}+t_{i}a_{2}+\epsilon_{i}$$

Where $y$ is the variable of interest (cognitive functioning) that changes with time $i$; $a_{1}$ and $a_{2}$ as the two latent variables representing the intercept and slope, respectively; $t_{i}$ as the value of time; and $\epsilon_{i}$ as the error term (i.e., how much individuals vary from their predicted trajectory).

Using the R (version 3.6.2) package ‘lavaan’, the following model can be specified for the intercept and slope within a SEM framework, where ‘cog_1’, ‘cog_2’ and ‘cog_3’ represent cognitive scores at each time period of assessment; ‘i’ represents the latent variable ‘cognition intercept’, and ‘s’ represents the latent variable ‘cognition slope’ (Fig. S2).

‘i =~ 1*cog_1 + 1*cog_2 + 1*cog_3

s =~ t_0_*cog_1 + t_1_*cog_2 + t_2_*cog_3’

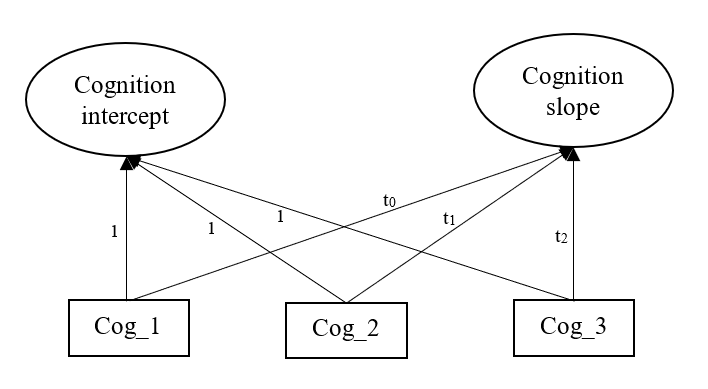

**Fig. S2**. A LGC model on cognitive functioning across 3 time points.

Coefficients t_0,_ t_1_ and t_2_ in the model will be calculated using the below formulae, where ‘Age’ represents the actual age of assessment for each participant at each follow-up period, which are subtracted by age at first assessment and divided by 10 to represent time in decades since the first assessment. These are averaged across the analytical sample to calculate the mean follow-up period from first assessment:

$$t_{0}=\Sigma\left\{ \frac{Age_{53}-Age_{53}}{10} \right\}/n$$

$$t_{1}=\Sigma\left\{ \frac{Age_{60-64}-Age_{53}}{10} \right\}/n$$

$$t_{2}=\Sigma\left\{ \frac{Age_{68-70}-Age_{53}}{10} \right\}/n$$

*Persistent financial adversity predicting change in cognitive functioning over time*

The impact of persistent financial adversity on cognition can be represented by the following formula, where $y$ represents the latent variables cognition intercept and slope, and $X$ represent the exposure variable (persistent financial adversity). This is then adjusted for the effect of covariates, denoted by $C$.

$$y=\beta_{0}+\beta_{1}X+\beta_{2}C+\epsilon$$

###### Effect modification model

The equation for the effect modification model is shown below, where $X$ represents the exposure variable persistent financial adversity; $M$ represents the moderator of interest (sex, childhood SES or APOE-ε4); $X^{*}M$ as the interaction term between the exposure and moderator; and $C$ as covariates in the model.

$$y=\beta_{0}+\beta_{1}X+\beta_{2}M+\beta_{3}X^{*}M+\beta_{4}C+\epsilon$$

##### ***Secondary analyses***

*Quantile regression*

The formula for a quantile regression model is shown below, where $Q$ is the conditional quantile of the outcome $y$ (i.e., cognitive decline), given a quantile level of $\tau$ which ranges between 0 and 1.

$Q_{\tau}\left( y \right)=\beta_{0}\left( \tau\right)+\beta_{1}\left( \tau\right)X+\epsilon$

Unlike linear regression, quantile regression offers insight into the effect of an exposure variable at different parts of the distributions of the outcome.^17^ Quantile regression was performed at the 25^th^ and 75^th^ quantile, representing those showing faster and slower decline, respectively.

##### ***Missing data***

###### Missing data due to attrition

To reduce potential drop-out bias, we calculated non-response weights using Inverse Probability Weighting. Predictors for non-response in the NSHD cohort have been reported previously.^1,18^ Multivariate regression model was used with these variables to predict likelihood of non-response, which were inverted to calculate non-response weightings. Missing data on predictor variables of non-response were imputed using multivariable imputation by chained equations (MICE),^19,20^ with 20 imputed datasets. Non-response weightings were pooled across the 20 datasets before merging with the analytical sample and used in subsequent analyses.

###### Missing data within the analytic sample

Auxiliary variables used for imputing missing data within the analytical sample included all variables in the analysis, along with other predictors of study variables, including personal income, occupational status, employment status, and marital status at 26, 36, 43 and 53 years. Imputation was performed on raw variables, with 20 imputed datasets generated, and any derived variables were calculated post imputation.

##### ***Sensitivity analyses***

The following four sensitivity analyses were performed:

Steps from primary analyses were repeated:

(1) Without the inclusion of sample weightings to examine whether findings were substantially different.
(2) Within Insight 46 to investigate to what extent findings from the main sample replicate within this sub-sample.

(3) Third, to check whether the classification of low household income may be affected by household composition, we calculated the bottom 20% of the sample within each family size grouping (0 to 5+) to derive a household size adjusted low income variable.

(4) Fourth, to check for potential implications of missing data strategies applied, we performed multiple imputation to handle missing data on cognitive function as well as all study variables prior to performing LGC analysis. The imputed datasets were then used to perform LGC analysis, first to estimate cognition intercept and slope, and second with added regression models to estimate the effect of financial adversity, with effects pooled across multiple imputed datasets.

##### ***Deviation from pre-registration***

We initially only proposed to test for the interaction between financial adversity and sex, childhood SEC, and APOE-ɛ4 on cognition slopes. However, we also tested these interactions on cognition intercepts (baseline at 53 years), given the associations observed with baseline scores.

***References***

1.	Kuh, D. *et al.* The MRC National Survey of Health and Development reaches age 70: maintaining participation at older ages in a birth cohort study. *Eur. J. Epidemiol.* **31**, 1135–1147 (2016).

2.	James, S.-N. *et al.* Using a birth cohort to study brain health and preclinical dementia: recruitment and participation rates in Insight 46. *BMC Res. Notes* **11**, 1–9 (2018).

3.	Office for National Statistics. Earnings timeseries of median gross weekly earnings from 1968 to 2021. https://www.ons.gov.uk/employmentandlabourmarket/peopleinwork/earningsandworkinghours/adhocs/13876earningstimeseriesofmediangrossweeklyearningsfrom1968to2021.

4.	Office for National Statistics. Average Incomes, Taxes and Benefits by Decile Groups of ALL Households. https://www.ons.gov.uk/peoplepopulationandcommunity/personalandhouseholdfinances/incomeandwealth/datasets/averageincomestaxesandbenefitsbydecilegroupsofallhouseholds.

### **Cognitive function outcomes**

**Table S1.** Regression models showing the association between indicators of financial adversity (tested for linear trends) and cognition at baseline (intercept) and decline over time (slope).

|  | β | 95%CI | p-value | β | 95%CI | p-value | β | 95%CI | p-value | β | 95%CI | p-value |
| --- | --- | --- | --- | --- | --- | --- | --- | --- | --- | --- | --- | --- |
|  | **Processing speed** | | | | | | **Verbal memory** | | | | | |
|  | **Baseline (intercept)** | | | **Decline (slope)** | | | **Baseline (intercept)** | | | **Decline (slope)** | | |
| **Low household income** |  |  |  |  |  |  |  |  |  |  |  |  |
| Unadjusted | **-0.13** | **(-0.18, -0.08)** | **0.000** | **0.01** | **(0.00, 0.01)** | **0.025** | **-0.34** | **(-0.40, -0.29)** | **0.000** | **0.02** | **(0.01, 0.02)** | **0.000** |
| Adjusted | **-0.07** | **(-0.13, -0.02)** | **0.004** | 0.01 | (-0.00, 0.01) | 0.099 | **-0.16** | **(-0.21, -0.11)** | **0.000** | **0.01** | **(0.00, 0.01)** | **0.000** |
| **Financial hardships** |  |  |  |  |  |  |  |  |  |  |  |  |
| Unadjusted | **-0.09** | **(-0.14, -0.04)** | **0.001** | 0.01 | (-0.00, 0.01) | 0.099 | **-0.24** | **(-0.30, -0.17)** | **0.000** | **0.01** | **(0.01, 0.01)** | **0.000** |
| Adjusted | **-0.05** | **(-0.11, -0.00)** | **0.040** | 0.00 | (-0.00, 0.01) | 0.219 | **-0.10** | **(-0.15, -0.05)** | **0.000** | **0.00** | **(0.00, 0.01)** | **0.023** |

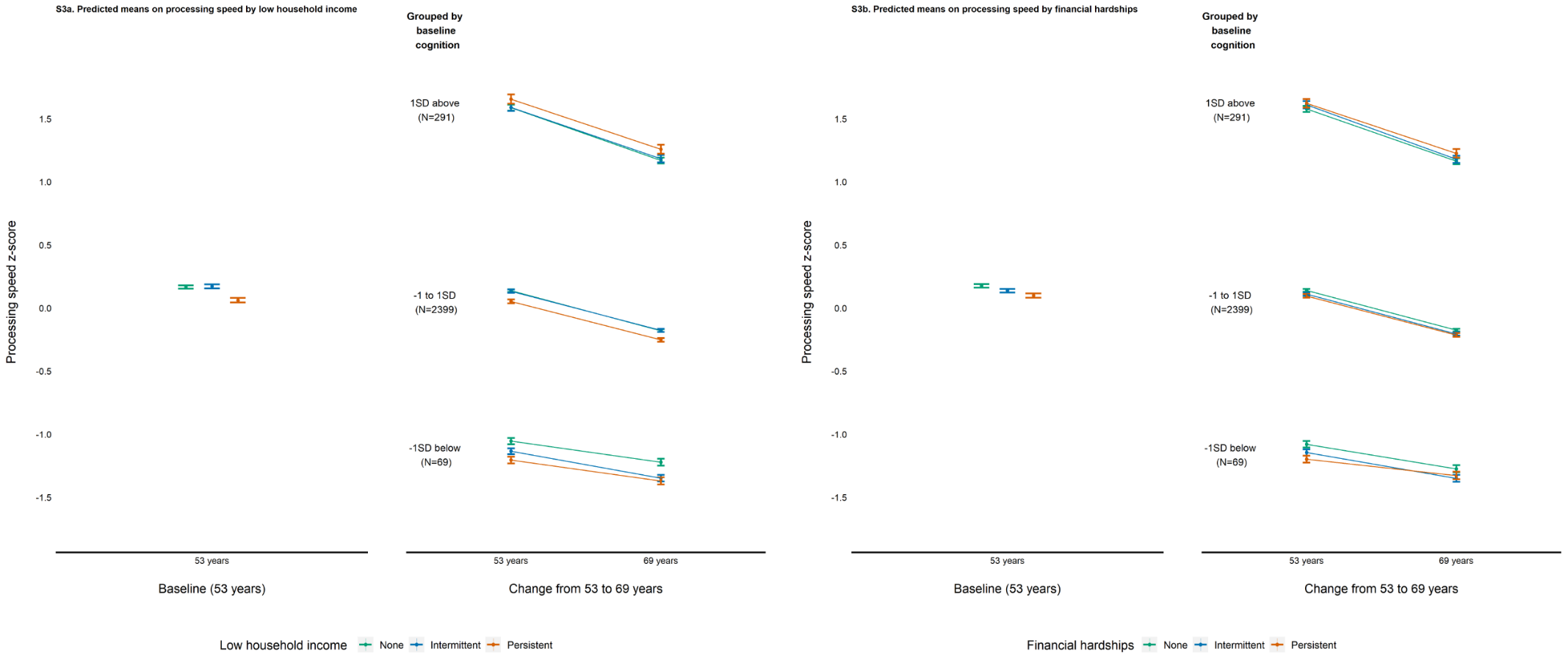

**Fig. S3**. Predicted means on processing speed at baseline (53 years) and decline (stratified on cognition at baseline) by financial adversity exposure.

**Table S2.** Regression models showing the association between indicators of financial adversity (coded as categorical variables) and cognitive decline.

|  | b | 95%CI | p.value | b | 95%CI | p.value |
| --- | --- | --- | --- | --- | --- | --- |
|  | **Processing speed (decline)** | | | **Verbal memory (decline)** | | |
| **Low household income** |  |  |  |  |  |  |
| Never [Reference] |  |  |  |  |  |  |
| Intermittent | 0.00 | (-0.01, 0.01) | 0.569 | 0.00 | (-0.00, 0.01) | 0.539 |
| Persistent | 0.01 | (-0.00, 0.02) | 0.099 | **0.01** | **(0.00, 0.01)** | **0.000** |
| **Financial hardships** |  |  |  |  |  |  |
| Never [Reference] |  |  |  |  |  |  |
| Intermittent | -0.00 | (-0.01, 0.01) | 0.831 | 0.00 | (-0.00, 0.01) | 0.364 |
| Persistent | 0.01 | (-0.00, 0.02) | 0.219 | **0.01** | **(0.00, 0.01)** | **0.023** |

All models adjusted for covariates.

**Table S3**. Regression models showing the association between indicators of financial adversity (as categorical variables) and cognitive decline at different quantiles, adjusted for all covariates.

|  | b | 95%CI | p.value | b | 95%CI | p.value |
| --- | --- | --- | --- | --- | --- | --- |
|  | **Processing speed (decline)** | | | **Verbal memory (decline)** | | |
| **Faster decline [25th quantile]** |  |  |  |  |  |  |
| Low household income [Intermittent] | 0.00 | (-0.01, 0.02) | 0.436 | -0.00 | (-0.01, 0.01) | 0.944 |
| Low household income [Persistent] | **0.02** | **(0.00, 0.03)** | **0.014** | **0.01** | **(0.00, 0.02)** | **0.002** |
| Financial hardships [Intermittent] | 0.00 | (-0.01, 0.01) | 0.664 | 0.00 | (-0.01, 0.01) | 0.624 |
| Financial hardships [Persistent] | 0.01 | (-0.01, 0.02) | 0.251 | **0.01** | **(0.00, 0.02)** | **0.014** |
| **Slower decline [75th quantile**] |  |  |  |  |  |  |
| Low household income [Intermittent] | 0.01 | (-0.01, 0.02) | 0.271 | 0.00 | (-0.00, 0.01) | 0.247 |
| Low household income [Persistent] | 0.00 | (-0.01, 0.01) | 0.686 | **0.01** | **(0.00, 0.02)** | **0.020** |
| Financial hardships [Intermittent] | 0.00 | (-0.01, 0.01) | 0.953 | 0.00 | (-0.00, 0.01) | 0.390 |
| Financial hardships [Persistent] | 0.00 | (-0.01, 0.02) | 0.564 | 0.01 | (-0.00, 0.01) | 0.211 |

When quantile regression was performed at the 25^th^ and 75^th^ quantile (faster and slower decline, respectively), only persistent experience of low household income was associated with verbal memory at both quantiles, and additionally associated with processing speed at the 25^th^ quantile only (faster decline) (supplementary materials Table S3).

**Table S4**. Regression models showing the interaction between indicators of financial adversity and sex, childhood SEC and APOE-4 on cognition at baseline (intercept) and decline (slope).

|  | b | 95%CI | p.value | b | 95%CI | p.value | b | 95%CI | p.value | b | 95%CI | p.value |
| --- | --- | --- | --- | --- | --- | --- | --- | --- | --- | --- | --- | --- |
|  | **Processing speed** | | |  | | | **Verbal memory** | | |  | | |
|  | **Baseline (intercept)** | | | **Decline (slope)** | | | **Baseline (intercept)** | | | **Decline (slope)** | | |
| **Low household income*** |  |  |  |  |  |  |  |  |  |  |  |  |
| Sex | **0.13** | **(0.03, 0.23)** | **0.009** | -0.00 | (-0.02, 0.01) | 0.721 | -0.04 | (-0.13, 0.05) | 0.402 | 0.00 | (-0.00, 0.01) | 0.525 |
| Childhood SEC | -0.08 | (-0.24, 0.07) | 0.308 | 0.01 | (-0.01, 0.03) | 0.430 | -0.02 | (-0.16, 0.13) | 0.810 | 0.00 | (-0.01, 0.01) | 0.515 |
| APOE-4 | 0.07 | (-0.05, 0.19) | 0.245 | 0.00 | (-0.01, 0.02) | 0.638 | -0.02 | (-0.13, 0.09) | 0.732 | -0.00 | (-0.01, 0.01) | 0.929 |
| **Financial hardships*** |  |  |  |  |  |  |  |  |  |  |  |  |
| Sex | 0.06 | (-0.04, 0.16) | 0.260 | **-0.01** | **(-0.03, 0.00)** | **0.075** | 0.00 | (-0.10, 0.10) | 0.994 | 0.01 | (-0.00, 0.01) | 0.131 |
| Childhood SEC | -0.08 | (-0.23, 0.07) | 0.294 | 0.01 | (-0.01, 0.03) | 0.283 | -0.08 | (-0.23, 0.06) | 0.259 | 0.00 | (-0.01, 0.01) | 0.833 |
| APOE-4 | 0.00 | (-0.12, 0.13) | 0.945 | -0.01 | (-0.03, 0.01) | 0.258 | 0.03 | (-0.09, 0.15) | 0.630 | -0.01 | (-0.01, 0.00) | 0.213 |

All models adjusted for covariates.

### **Brain health outcomes**

**Table S5.** Regression models showing the association between indicators of financial adversity (tested for linear trends) and β-amyloid (Aβ) and baseline volumetric measures.

|  | **Main effect of financial adversity**^a^ | | | **Main effect of moderators**^b^ | | | **Interactions**^b^ | | |
| --- | --- | --- | --- | --- | --- | --- | --- | --- | --- |
|  | OR | 95%CI | p.value | OR | 95%CI | p.value | OR | 95%CI | p.value |
|  | **Aβ status** | | |  |  |  |  |  |  |
| **Low household income** | 0.02 | (-0.69, 0.73) | 0.960 |  |  |  |  |  |  |
| **Sex** |  |  |  | -0.03 | (-0.59, 0.53) | 0.909 | -0.44 | (-1.89, 1.02) | 0.556 |
| **Child SEC** |  |  |  | 0.74 | (-0.33, 1.81) | 0.175 | 0.70 | (-1.27, 2.66) | 0.485 |
| **APOE-ɛ4** |  |  |  | **1.65** | **(1.09, 2.22)** | **<0.001** | -0.16 | (-1.87, 1.54) | 0.850 |
| **Financial hardships** | 0.15 | (-0.37, 0.68) | 0.566 |  |  |  |  |  |  |
| **Sex** |  |  |  | 0.04 | (-0.53, 0.61) | 0.889 | 0.39 | (-0.71, 1.49) | 0.486 |
| **Child SEC** |  |  |  | 0.72 | (-0.33, 1.77) | 0.176 | -0.57 | (-2.29, 1.15) | 0.516 |
| **APOE-ɛ4** |  |  |  | **1.58** | **(1.02, 2.13)** | **<0.001** | 0.38 | (-0.79, 1.55) | 0.522 |
|  | B | 95%CI | p.value | B | 95%CI | p.value | B | 95%CI | p.value |
|  | **Aβ burden (SUVR)** | | |  |  |  |  |  |  |
| **Low household income** | 0.00 | (-0.01, 0.02) | 0.602 |  |  |  |  |  |  |
| **Sex** |  |  |  | -0.00 | (-0.02, 0.01) | 0.726 | -0.02 | (-0.06, 0.02) | 0.313 |
| **Child SEC** |  |  |  | 0.01 | (-0.02, 0.04) | 0.349 | 0.03 | (-0.03, 0.10) | 0.337 |
| **APOE-ɛ4** |  |  |  | **0.06** | **(0.04, 0.07)** | **<0.001** | 0.01 | (-0.04, 0.06) | 0.764 |
| **Financial hardships** | 0.01 | (-0.01, 0.02) | 0.401 |  |  |  |  |  |  |
| **Sex** |  |  |  | -0.00 | (-0.02, 0.01) | 0.790 | 0.02 | (-0.01, 0.05) | 0.170 |
| **Child SEC** |  |  |  | 0.01 | (-0.01, 0.04) | 0.314 | 0.02 | (-0.05, 0.08) | 0.606 |
| **APOE-ɛ4** |  |  |  | **0.05** | **(0.04, 0.07)** | **<0.001** | **0.03** | **(-0.01, 0.06)** | **0.099** |
|  | **WMHV**^c^ | | |  |  |  |  |  |  |
| **Low household income** | 0.01 | (-0.24, 0.27) | 0.918 |  |  |  |  |  |  |
| **Sex** |  |  |  | **0.35** | **(0.10, 0.60)** | **0.007** | 0.07 | (-0.45, 0.58) | 0.804 |
| **Child SEC** |  |  |  | 0.18 | (-0.24, 0.59) | 0.406 | -0.08 | (-0.86, 0.71) | 0.851 |
| **APOE-ɛ4** |  |  |  | 0.11 | (-0.11, 0.33) | 0.332 | -0.06 | (-0.84, 0.72) | 0.874 |
| **Financial hardships** | 0.08 | (-0.13, 0.29) | 0.437 |  |  |  |  |  |  |
| **Sex** |  |  |  | **0.31** | **(0.06, 0.57)** | **0.016** | 0.07 | (-0.36, 0.50) | 0.745 |
| **Child SEC** |  |  |  | 0.16 | (-0.25, 0.57) | 0.445 | -0.24 | (-1.04, 0.57) | 0.563 |
| **APOE-ɛ4** |  |  |  | 0.12 | (-0.11, 0.34) | 0.307 | 0.03 | (-0.44, 0.49) | 0.911 |
|  | **Baseline total brain volume** | | |  |  |  |  |  |  |
| **Low household income** | -4.20 | (-15.25, 6.89) | 0.455 |  |  |  |  |  |  |
| **Sex** |  |  |  | **22.59** | **(11.45, 33.73)** | **<0.001** | 5.34 | (-17.12, 27.80) | 0.640 |
| **Child SEC** |  |  |  | 5.14 | (-11.71, 21.99) | 0.549 | 8.65 | (-24.71, 42.00) | 0.610 |
| **APOE-ɛ4** |  |  |  | **9.91** | **(0.14, 19.68)** | **0.047** | -16.74 | (-45.43, 11.94) | 0.252 |
| **Financial hardships** | 6.79 | (-2.36, 15.95) | 0.146 |  |  |  |  |  |  |
| **Sex** |  |  |  | **19.35** | **(8.22, 30.47)** | **0.001** | 10.80 | (-7.91, 29.51) | 0.257 |
| **Child SEC** |  |  |  | 3.51 | (-12.99, 20.01) | 0.676 | -16.13 | (-51.56, 19.30) | 0.370 |
| **APOE-ɛ4** |  |  |  | **10.58** | **(1.01, 20.16)** | **0.030** | -2.61 | (-22.62, 17.39) | 0.797 |
|  | **Baseline hippocampal volume** | | |  |  |  |  |  |  |
| **Low household income** | -0.01 | (-0.16, 0.14) | 0.894 |  |  |  |  |  |  |
| **Sex** |  |  |  | -0.11 | (-0.26, 0.04) | 0.136 | 0.18 | (-0.14, 0.50) | 0.272 |
| **Child SEC** |  |  |  | -0.17 | (-0.41, 0.06) | 0.137 | -0.07 | (-0.56, 0.42) | 0.766 |
| **APOE-ɛ4** |  |  |  | -0.06 | (-0.19, 0.06) | 0.316 | 0.12 | (-0.29, 0.53) | 0.559 |
| **Financial hardships** | 0.07 | (-0.05, 0.19) | 0.270 |  |  |  |  |  |  |
| **Sex** |  |  |  | -0.14 | (-0.28, 0.01) | 0.067 | 0.11 | (-0.14, 0.37) | 0.372 |
| **Child SEC** |  |  |  | -0.19 | (-0.41, 0.04) | 0.108 | -0.02 | (-0.47, 0.44) | 0.943 |
| **APOE-ɛ4** |  |  |  | -0.06 | (-0.19, 0.06) | 0.337 | -0.06 | (-0.34, 0.21) | 0.648 |
|  | **Baseline ventricular volume** | | |  |  |  |  |  |  |
| **Low household income** | **4.67** | **(1.01, 8.32)** | **0.013** |  |  |  |  |  |  |
| **Sex** |  |  |  | 1.14 | (-2.41, 4.68) | 0.530 | -5.29 | (-12.60, 2.02) | 0.155 |
| **Child SEC** |  |  |  | -1.40 | (-6.68, 3.89) | 0.604 | 4.82 | (-6.64, 16.28) | 0.408 |
| **APOE-ɛ4** |  |  |  | -1.43 | (-4.52, 1.66) | 0.363 | 3.72 | (-6.08, 13.53) | 0.455 |
| **Financial hardships** | 0.10 | (-2.90, 3.10) | 0.946 |  |  |  |  |  |  |
| **Sex** |  |  |  | 1.77 | (-1.85, 5.39) | 0.338 | **-6.20** | **(-12.30, -0.10)** | **0.046** |
| **Child SEC** |  |  |  | -7.22 | (-6.08, 4.63) | 0.791 | 0.13 | (-11.25, 11.51) | 0.982 |
| **APOE-ɛ4** |  |  |  | -1.72 | (-4.7, 1.32) | 0.266 | 3.04 | (-3.67, 9.74) | 0.373 |

^a^Adjusted for sex and intracranial volume (for WMHV and baseline volumetric measures)

^b^Adjusted for all covariates

^c^Log-transformed

**Table S6**. Regression models showing the association between indicators of financial adversity (tested for linear trends) and longitudinal changes in brain health (per year over 2.4 years).

|  | **Main effect of financial adversity**^a^ | | | **Main effect of moderators**^b^ | | | **Interactions**^b^ | | |
| --- | --- | --- | --- | --- | --- | --- | --- | --- | --- |
|  | B | 95%CI | p.value | B | 95%CI | p.value | B | 95%CI | p.value |
|  | **Total brain volume atrophy** | | |  | | |  | | |
| **Low household income** | -0.20 | (-1.19, 0.79) | 0.696 |  |  |  |  |  |  |
| **Sex** |  |  |  | 0.59 | (-0.36, 1.54) | 0.220 | -0.70 | (-2.68, 1.27) | 0.483 |
| **Child SEC** |  |  |  | 0.00 | (-1.45, 1.45) | 0.998 | 1.58 | (-2.55, 5.71) | 0.446 |
| **APOE-ɛ4** |  |  |  | **1.06** | **(0.20, 1.93)** | **0.016** | **3.96** | **(0.53, 7.39)** | **0.025** |
| **Financial hardships** | 0.42 | (-0.38, 1.22) | 0.302 |  |  |  |  |  |  |
| **Sex** |  |  |  | 0.41 | (-0.54, 1.36) | 0.398 | **-2.37** | **(-3.95, -0.80)** | **0.003** |
| **Child SEC** |  |  |  | -0.04 | (-1.50, 1.41) | 0.953 | 1.40 | (-2.48, 5.29) | 0.473 |
| **APOE-ɛ4** |  |  |  | **1.10** | **(0.24, 1.95)** | **0.012** | **1.63** | **(-0.06, 3.31)** | **0.058** |
|  | **Total hippocampal volume atrophy** | | |  |  |  |  |  |  |
| **Low household income** | 0.00 | (-0.01, 0.01) | 0.908 |  |  |  |  |  |  |
| **Sex** |  |  |  | 0.01 | (-0.01, 0.02) | 0.376 | -0.01 | (-0.04, 0.02) | 0.506 |
| **Child SEC** |  |  |  | 0.00 | (-0.02, 0.02) | 0.852 | **0.06** | **(0.01, 0.12)** | **0.033** |
| **APOE-ɛ4** |  |  |  | **0.01** | **(0.00, 0.02)** | **0.024** | **0.06** | **(0.01, 0.10)** | **0.028** |
| **Financial hardships** | 0.01 | (-0.00, 0.02) | 0.169 |  |  |  |  |  |  |
| **Sex** |  |  |  | 0.00 | (-0.01, 0.02) | 0.699 | -0.01 | (-0.03, 0.01) | 0.524 |
| **Child SEC** |  |  |  | 0.00 | (-0.02, 0.02) | 0.871 | 0.04 | (-0.02, 0.11) | 0.197 |
| **APOE-ɛ4** |  |  |  | **0.01** | **(0.00, 0.02)** | **0.021** | **0.03** | **(0.01, 0.06)** | **0.004** |
|  | **Ventricular expansion** | | |  |  |  |  |  |  |
| **Low household income** | 0.21 | (-0.10, 0.52) | 0.188 |  |  |  |  |  |  |
| **Sex** |  |  |  | 0.18 | (-0.13, 0.48) | 0.256 | -0.09 | (-0.72, 0.53) | 0.767 |
| **Child SEC** |  |  |  | 0.36 | (-0.09, 0.81) | 0.120 | **1.42** | **(0.14, 2.69)** | **0.031** |
| **APOE-ɛ4** |  |  |  | **0.33** | **(0.07, 0.59)** | **0.013** | **1.66** | **(0.51, 2.82)** | **0.006** |
| **Financial hardships** | 0.21 | (-0.04, 0.46) | 0.105 |  |  |  |  |  |  |
| **Sex** |  |  |  | 0.14 | (-0.17, 0.45) | 0.379 | **-0.73** | **(-1.24, -0.22)** | **0.005** |
| **Child SEC** |  |  |  | 0.37 | (-0.08, 0.82) | 0.111 | 1.34 | (-0.38, 3.06) | 0.122 |
| **APOE-ɛ4** |  |  |  | **0.32** | **(0.06, 0.58)** | **0.017** | **0.83** | **(0.30, 1.36)** | **0.002** |

^a^Adjusted for sex and intracranial volume

^b^Adjusted for all covariates

### **Additional analyses**

We additionally tested for an interaction between persistent experience of financial adversity and sex, childhood SEC and APOE-ɛ4 on verbal memory decline only, given that only persistent experience, rather than intermittent experience, was associated with verbal memory decline from secondary analyses (supplementary materials Table S2). No evidence of interactions was found (supplementary materials Table S6). Furthermore, since both low household income and financial hardships were also associated with processing speed (baseline) and verbal memory (baseline and decline) from primary analyses, we further included an interaction term between them, and no interactions were found (supplementary materials Table S7).

**Table S7**. Regression models showing the interaction between indicators of financial adversity and sex, childhood SEC and APOE-4 in relation to verbal memory decline.

|  | b | 95%CI | p.value | b | 95%CI | p.value |
| --- | --- | --- | --- | --- | --- | --- |
| **Verbal memory decline** | | | | | | |
|  | **Persistent low household income** | | | **Persistent financial hardships** | | |
| **Interaction with** |  |  |  |  |  |  |
| Sex | 0.00 | (-0.00, 0.01) | 0.325 | 0.01 | (-0.00, 0.02) | 0.175 |
| Childhood SEC | 0.00 | (-0.01, 0.02) | 0.828 | 0.00 | (-0.01, 0.01) | 0.980 |
| APOE-4 | -0.00 | (-0.01, 0.01) | 0.941 | -0.01 | (-0.02, 0.00) | 0.153 |

**Table S8**. Regression models showing the interaction between low household income and financial hardships on verbal memory.

|  | b | 95%CI | p.value | b | 95%CI | p.value | b | 95%CI | p.value |
| --- | --- | --- | --- | --- | --- | --- | --- | --- | --- |
|  | **Processing speed (intercept)** | | | **Verbal memory (intercept)** | | | **Verbal memory (decline)** | | |
| **Low household income x Financial hardships** | -0.08 | (-0.17, 0.02) | 0.113 | -0.00 | (-0.10, 0.09) | 0.951 | 0.00 | (-0.01, 0.01) | 0.811 |

### **Sensitivity analyses**

#### 1. **Repeat primary analyses without sample weighting**

**Table S9**. Regression models showing the association between indicators of financial adversity (tested for linear trends) and cognition at endpoint (intercept) and decline over time (slope), without sample weights.

|  | b | 95%CI | p.value | b | 95%CI | p.value | b | 95%CI | p.value | b | 95%CI | p.value |
| --- | --- | --- | --- | --- | --- | --- | --- | --- | --- | --- | --- | --- |
|  | **Processing speed** | | | | | | **Verbal memory** | | | | | |
|  | **Endpoint (intercept)** | | | **Decline (slope)** | | | **Endpoint (intercept)** | | | **Decline (slope)** | | |
| **Low household income** |  |  |  |  |  |  |  |  |  |  |  |  |
| Unadjusted | **-0.13** | **(-0.18, -0.08)** | **0.000** | **0.01** | **(0.00, 0.02)** | **0.033** | **-0.34** | **(-0.40, -0.29)** | **0.000** | **0.01** | **(0.01, 0.01)** | **0.000** |
| Adjusted | **-0.07** | **(-0.13, -0.02)** | **0.006** | 0.01 | (-0.00, 0.01) | 0.095 | **-0.16** | **(-0.21, -0.11)** | **0.000** | **0.00** | **(0.00, 0.01)** | **0.000** |
| **Financial hardships** |  |  |  |  |  |  |  |  |  |  |  |  |
| Unadjusted | **-0.08** | **(-0.14, -0.02)** | **0.011** | 0.00 | (-0.00, 0.01) | 0.237 | **-0.21** | **(-0.27, -0.15)** | **0.000** | **0.01** | **(0.00, 0.01)** | **0.000** |
| Adjusted | -0.04 | (-0.10, 0.02) | 0.162 | 0.00 | (-0.00, 0.01) | 0.396 | **-0.09** | **(-0.14, -0.04)** | **0.000** | **0.00** | **(0.00, 0.00)** | **0.024** |

**Table S10.** Regression models showing the interaction between indicators of financial adversity and sex, childhood SEC and APOE-4 in relation to cognitive decline, adjusted for all covariates, without sample weights.

|  | **b** | **95%CI** | **p.value** | **b** | **95%CI** | **p.value** |
| --- | --- | --- | --- | --- | --- | --- |
|  | **Processing speed (decline)** | | | **Verbal memory (decline)** | | |
| **Low household income*** |  |  |  |  |  |  |
| Sex | -0.00 | (-0.01, 0.01) | 0.965 | 0.00 | (-0.00, 0.00) | 0.690 |
| Childhood SEC | 0.01 | (-0.01, 0.03) | 0.308 | 0.00 | (-0.00, 0.01) | 0.529 |
| APOE-4 | 0.00 | (-0.01, 0.02) | 0.611 | 0.00 | (-0.00, 0.00) | 0.911 |
| **Financial hardships*** |  |  |  |  |  |  |
| Sex | -0.01 | (-0.03, 0.00) | 0.165 | 0.00 | (-0.00, 0.01) | 0.425 |
| Childhood SEC | 0.01 | (-0.01, 0.04) | 0.311 | 0.00 | (-0.00, 0.01) | 0.606 |
| APOE-4 | -0.01 | (-0.03, 0.01) | 0.356 | -0.00 | (-0.01, 0.00) | 0.293 |

#### 2. **Repeat primary analyses in Insight46 sub-sample**

**Table S11.** Regression models showing the association between indicators of financial adversity (tested for linear trends) and cognition at endpoint (intercept) and decline over time (slope), in Insight46 sub-sample only.

|  | b | 95%CI | p.value | b | 95%CI | p.value | b | 95%CI | p.value | b | 95%CI | p.value |
| --- | --- | --- | --- | --- | --- | --- | --- | --- | --- | --- | --- | --- |
|  | **Processing speed** | | | | | | **Verbal memory** | | | | | |
|  | **Endpoint (intercept)** | | | **Decline (slope)** | | | **Endpoint (intercept)** | | | **Decline (slope)** | | |
| **Low household income** |  |  |  |  |  |  |  |  |  |  |  |  |
| Unadjusted | -0.06 | (-0.21, 0.09) | 0.422 | 0.02 | (-0.00, 0.05) | 0.104 | **-0.29** | **(-0.46, -0.12)** | **0.001** | **0.02** | **(0.00, 0.03)** | **0.017** |
| Adjusted | -0.05 | (-0.21, 0.10) | 0.516 | 0.02 | (-0.01, 0.05) | 0.126 | **-0.25** | **(-0.40, -0.10)** | **0.001** | **0.01** | **(0.00, 0.03)** | **0.026** |
| **Financial hardships** |  |  |  |  |  |  |  |  |  |  |  |  |
| Unadjusted | -0.05 | (-0.17, 0.08) | 0.447 | 0.01 | (-0.01, 0.03) | 0.242 | -0.03 | (-0.18, 0.11) | 0.647 | -0.00 | (-0.01, 0.01) | 0.414 |
| Adjusted | -0.06 | (-0.19, 0.06) | 0.331 | 0.01 | (-0.01, 0.03) | 0.231 | -0.06 | (-0.18, 0.07) | 0.378 | -0.00 | (-0.01, 0.01) | 0.483 |

**Table S12**. Regression models showing the interaction between indicators of financial adversity and sex, childhood SEC and APOE-4 in relation to cognitive decline, adjusted for all covariates, in Insight46 sub-sample only.

|  | **b** | **95%CI** | **p.value** | **b** | **95%CI** | **p.value** |
| --- | --- | --- | --- | --- | --- | --- |
|  | **Processing speed (decline)** | | | **Verbal memory (decline)** | | |
| **Low household income*** |  |  |  |  |  |  |
| Sex | 0.02 | (-0.04, 0.07) | 0.537 | 0.00 | (-0.02, 0.03) | 0.761 |
| Childhood SEC | -0.01 | (-0.08, 0.07) | 0.827 | -0.01 | (-0.04, 0.03) | 0.667 |
| APOE-4 | 0.05 | (-0.02, 0.13) | 0.160 | -0.01 | (-0.04, 0.03) | 0.739 |
| **Financial hardships*** |  |  |  |  |  |  |
| Sex | -0.02 | (-0.06, 0.03) | 0.483 | **0.03** | **(0.01, 0.05)** | **0.008** |
| Childhood SEC | 0.02 | (-0.06, 0.09) | 0.700 | 0.01 | (-0.03, 0.05) | 0.719 |
| APOE-4 | -0.03 | (-0.08, 0.02) | 0.219 | 0.01 | (-0.01, 0.03) | 0.258 |

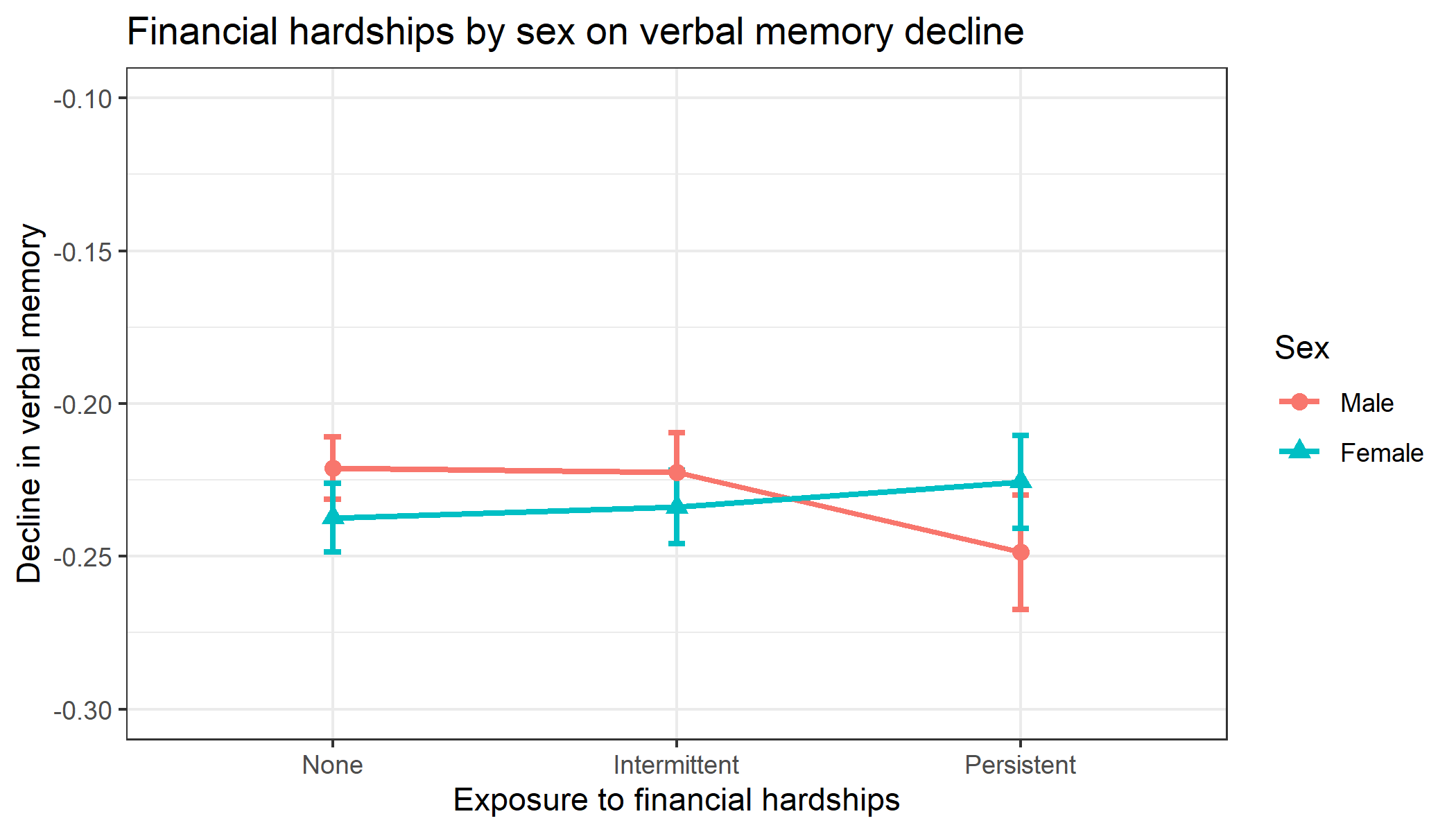

**Fig. S4**. Insight46 sub-sample: Predicted means on verbal memory decline by financial hardships exposure and sex.

#### 3. **Repeat primary analyses when low household income is derived within each family size grouping**

**Table S13**. Regression models showing the association between indicators of financial adversity and cognition at endpoint (intercept) and decline over time (slope).

|  | b | 95%CI | p.value | b | 95%CI | p.value | b | 95%CI | p.value | b | 95%CI | p.value |
| --- | --- | --- | --- | --- | --- | --- | --- | --- | --- | --- | --- | --- |
|  | **Processing speed** | | | | | | **Verbal memory** | | | | | |
|  | **Endpoint (intercept)** | | | **Decline (slope)** | | | **Endpoint (intercept)** | | | **Decline (slope)** | | |
| **Low household income**^a^ |  |  |  |  |  |  |  |  |  |  |  |  |
| Unadjusted | **-0.10** | **(-0.15, -0.06)** | **0.000** | **0.01** | **(0.00, 0.02)** | **0.022** | **-0.35** | **(-0.40, -0.30)** | **0.000** | **0.01** | **(0.01, 0.02)** | **0.000** |
| Adjusted | -0.04 | (-0.10, 0.01) | 0.081 | 0.01 | (-0.00, 0.01) | 0.118 | **-0.16** | **(-0.20, -0.11)** | **0.000** | **0.00** | **(0.00, 0.01)** | **0.004** |
| **Financial hardships** |  |  |  |  |  |  |  |  |  |  |  |  |
| Unadjusted | **-0.09** | **(-0.14, -0.03)** | **0.001** | 0.01 | (-0.00, 0.01) | 0.101 | **-0.23** | **(-0.29, -0.17)** | **0.000** | **0.01** | **(0.01, 0.01)** | **0.000** |
| Adjusted | -0.05 | (-0.10, 0.00) | 0.054 | 0.00 | (-0.00, 0.01) | 0.223 | **-0.10** | **(-0.15, -0.05)** | **0.000** | **0.00** | **(0.00, 0.01)** | **0.020** |

^a^ Low household income is adjusted for household size

**Table S14**. Regression models showing the interaction between indicators of financial adversity and sex, childhood SEC and APOE-4 in relation to cognitive decline, adjusted for all covariates (with low household income adjusted for household size).

|  | **b** | **95%CI** | **p.value** | **b** | **95%CI** | **p.value** |
| --- | --- | --- | --- | --- | --- | --- |
|  | **Processing speed (decline)** | | | **Verbal memory (decline)** | | |
| **Low household income*** |  |  |  |  |  |  |
| Sex | -0.00 | (-0.02, 0.01) | 0.646 | 0.00 | (-0.00, 0.01) | 0.489 |
| Childhood SEC | -0.00 | (-0.03, 0.02) | 0.708 | 0.00 | (-0.01, 0.02) | 0.400 |
| APOE-4 | 0.01 | (-0.01, 0.02) | 0.314 | 0.00 | (-0.01, 0.01) | 0.998 |
| **Financial hardships*** |  |  |  |  |  |  |
| Sex | **-0.01** | **(-0.03, 0.00)** | **0.060** | 0.01 | (-0.00, 0.01) | 0.144 |
| Childhood SEC | 0.01 | (-0.01, 0.04) | 0.259 | 0.00 | (-0.01, 0.01) | 0.826 |
| APOE-4 | -0.01 | (-0.03, 0.01) | 0.265 | -0.01 | (-0.01, 0.00) | 0.246 |

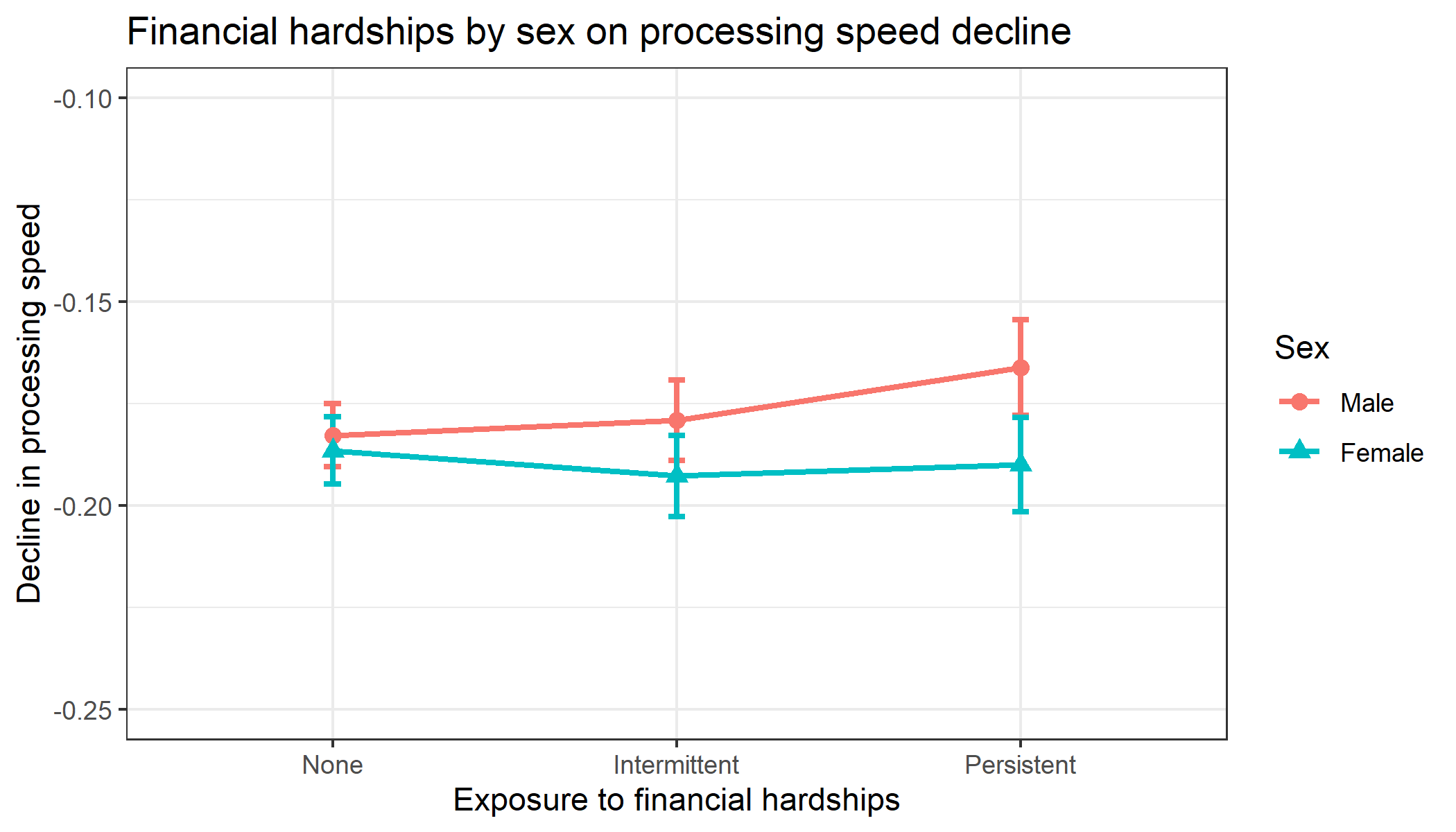

**Fig. S5**. Low household income by household size: Predicted means on processing speed decline by financial hardships exposure and sex.

#### 4. **Perform multiple imputation on cognitive scores and estimate cognitive decline and the role of financial adversity within SEM framework**

**Table S15.** Standardised estimates showing the association between financial adversity (low household income and financial hardships) and processing speed and verbal memory (intercept and slope).

|  | b | 95%CI | p.value | b | 95%CI | p.value |
| --- | --- | --- | --- | --- | --- | --- |
|  | **Unadjusted** |  |  | **Adjusted** |  |  |
| **Processing speed** |  |  |  |  |  |  |
| Intercept ~ low household income | **-0.13** | **(-0.19, -0.07)** | **0.000** | **-0.08** | **(-0.14, -0.02)** | **0.012** |
| Slope ~ low household income | 0.03 | (-0.01, 0.06) | 0.128 | 0.03 | (-0.01, 0.06) | 0.154 |
| Intercept ~ financial hardships | **-0.08** | **(-0.15, -0.02)** | **0.009** | -0.05 | (-0.11, 0.01) | 0.104 |
| Slope ~ financial hardships | 0.01 | (-0.03, 0.04) | 0.766 | 0.00 | (-0.03, 0.04) | 0.857 |
| **Verbal memory** |  |  |  |  |  |  |
| Intercept ~ low household income | **-0.30** | **(-0.35, -0.24)** | **0.000** | **-0.13** | **(-0.18, -0.08)** | **0.000** |
| Slope ~ low household income | 0.02 | (-0.01, 0.05) | 0.269 | -0.00 | (-0.03, 0.03) | 0.886 |
| Intercept ~ financial hardships | **-0.20** | **(-0.26, -0.14)** | **0.000** | **-0.09** | **(-0.14, -0.04)** | **0.001** |
| Slope ~ financial hardships | 0.01 | (-0.02, 0.04) | 0.579 | -0.00 | (-0.03, 0.03) | 0.801 |
